# Home birth and associated factors in Nigeria: a comparative study of rural and urban settings based on the analysis of national population-based data

**DOI:** 10.1101/2025.05.31.25328693

**Authors:** Emmanuel O Adewuyi, Asa Auta, Olumuyiwa Omonaiye, Mary I Adewuyi, Victory Olutuase, Kazeem Adefemi, Olumide Odeyemi, Yun Zhao, Gizachew A Tessema, Gavin Pereira

## Abstract

**Introduction:** Despite global efforts to reduce maternal and neonatal mortality, Nigeria continues to report disproportionately high rates. Home birth, childbirth occurring outside health facilities and without timely access to emergency obstetric care, remains a significant public health concern. National estimates can obscure stark sub-national disparities. This study estimated the prevalence of home birth and identified associated factors, comparing national, rural, and urban contexts.

**Methods:** We analysed data from 21,512 mothers using the 2018 Nigeria Demographic and Health Survey, guided by Andersen’s Behavioural Model. Logistic regression was used to examine associations between home birth and a range of individual, household, and contextual factors.

**Results:** Nationally, 58.1% (95% CI: 56.5, 59.7) of mothers gave birth at home, with prevalence nearly twice as high in rural areas (72.4%) compared to urban areas (36.1%). The North-West region reported the highest prevalence nationally (83.6%), in rural (89.4%) and urban (66.6%) areas. The South-East had the lowest prevalence in rural areas (16.2%), and the South-West in urban areas (16.7%). Nationally and across settings, lower maternal and partner education, poor household wealth, fewer than eight antenatal contacts, higher birth order, Hausa-Fulani ethnicity, and limited exposure to media and the internet were associated with increased odds of home birth. In rural areas, additional predictors included greater difficulty obtaining permission to seek care, distance to facilities, limited maternal decision-making autonomy, and pronounced regional disparities across all northern and the South-South regions. In urban areas, younger maternal age, Islamic religion, and financial barriers to accessing healthcare were uniquely associated.

**Conclusion:** Home birth remains common in Nigeria, particularly in rural areas and in northern and South-South regions, reflecting persistent structural, socioeconomic, and informational inequities. Addressing home birth requires setting-specific, equity-oriented strategies. In rural areas, policies should prioritise women’s autonomy, reduce geographic and regional barriers, and expand healthcare access. In urban areas, targeted interventions should focus on supporting younger mothers, mitigating financial barriers, and providing culturally and religiously responsive care. Nationally, investments in education, antenatal care utilisation, and access to health information through media and the internet are critical to promoting facility-based childbirth and improving maternal and neonatal outcomes.

## Introduction

Reducing maternal and neonatal mortality remains a global public health priority, with initiatives such as the Millennium Development Goals (MDGs) and Sustainable Development Goals (SDGs) contributing to notable progress [1, 2]. Maternal mortality refers to the death of a woman from pregnancy-related causes up to 42 days postpartum, while neonatal mortality refers to the death of an infant within the first 28 days of life [3, 4]. Despite the global gains, many low-resource settings continue to experience disproportionately high mortality rates, largely due to persistent inequities in the availability, accessibility, and utilisation of quality healthcare services [2, 5, 6]. Nigeria exemplifies this ongoing challenge: the country accounts for over 28% of global maternal deaths and ranks second in the absolute number of neonatal deaths [3, 4, 7]. The World Health Organisation (WHO) data report a maternal mortality ratio of 1,047 deaths per 100,000 live births, classified as extremely high [3, 4]. The Nigerian government has introduced initiatives such as the Midwives Service Scheme and conditional cash transfers to improve access and service utilisation [8–11]. Nonetheless, maternal and neonatal mortality remain unacceptably high in Nigeria, with the continued prevalence of home births emerging as a likely key contributing factor.

Home births, defined as childbirths occurring outside health facilities without skilled attendance (e.g., obstetricians or midwives) and timely access to emergency care, are linked with increased maternal and neonatal risks [12–14]. The lack of emergency obstetric care in these settings often leads to preventable complications and deaths [12]. In contrast, health facility births attended by skilled personnel offer better outcomes through timely management of complications such as severe bleeding, infections, obstructed labour, and eclampsia—major causes of maternal and neonatal mortality [12, 15–18]. These complications are often unpredictable but treatable in facilities equipped and staffed for emergency obstetric care services [12, 14]. Home birth settings in Nigeria and many low- to middle-income countries typically lack resources for emergency care or timely health facility transfer, heightening the risk of adverse outcomes [17]. Notably, most maternal deaths occur during labour, childbirth, or the immediate postpartum period—the most critical window for survival [3, 4, 15]. Hence, ensuring facility-based births with access to emergency care remains essential to reducing these deaths.

Despite the well-established benefits of health facility childbirth, home births remain common in Nigeria [12, 18–23]. Understanding the factors associated with this practice is crucial for developing effective, targeted interventions. Prior research suggests that low maternal education, limited household wealth, and logistical barriers are among the significant factors associated with maternal health services utilisation [19–22, 24–26]. However, to our knowledge, most studies on this subject in Nigeria [19–22, 26] rely on pooled national data, which may obscure important subnational differences [25]. Increasingly, the value of disaggregated data is recognised in identifying inequities across geographic and socioeconomic boundaries [27, 28]. For example, the WHO’s framework for monitoring progress toward Universal Health Coverage (UHC) underscores the importance of disaggregated data in assessing equity in healthcare access [29]. Similarly, the 2024 Operational Framework for Monitoring Social Determinants of Health Equity highlights how disaggregation by geographic location (e.g., rural versus urban), demographic factors (e.g., age and sex), and socioeconomic status can reveal disparities and guide targeted interventions [30]. These approaches are central to achieving the 2030 Agenda for Sustainable Development, which aims to ‘leave no one behind’ [30].

Nigeria serves as a compelling setting for the use of disaggregated data due to its highly diverse population [13]. Geographical, ethnic, demographic, cultural, and socioeconomic heterogeneity in the country compounds existing health inequities, creating critical implications for policymaking and resource allocation. Accordingly, understanding home birth patterns across subpopulations is essential for addressing health inequities and improving outcomes. Tailored strategies based on such insights can target areas of greatest need. Aligning with this premise, a previous study assessed the prevalence and factors associated with the non-utilisation of healthcare facilities for childbirth in Nigeria, focusing on differences between rural and urban populations [19]. Given the complexity of the issue and the need for more granular insights, the present study builds on those findings by further examining home birth patterns using the most recent nationally representative demographic and health survey data. Unlike the previous study [19], this research estimates home birth prevalence and assesses associated factors both nationally and across rural and urban areas, and, to our knowledge, is the first in Nigeria to do so.

To achieve our study objectives, we employed Andersen’s Behavioural Model of Healthcare Utilisation [31], a theory-driven framework that systematically examines how sociodemographic, economic, and health system factors influence healthcare use, in this case, home birth. We also applied a social determinants of health lens, which considers the broader context in which people are born, live, work, and age, along with a social justice perspective that highlights structural inequities, in the discussion of our findings [30, 32–34]. This dual lens shifts the focus beyond individual behaviours to systemic constraints in maternal healthcare access. Further, our study offers policy recommendations aimed at empowering women, reducing home births and, subsequently, improving maternal and neonatal outcomes. Findings are expected to contribute to achieving SDG targets 3.1 (reducing maternal mortality to <70 per 100,000 live births) and 3.2 (ending preventable deaths of newborns and children under five) by 2030 [1, 2], in Nigeria.

## Methods

### Study Setting

Nigeria is in West Africa, between 4° and 14° N latitude and 3° and 14° E longitude. It borders Benin, Niger, Chad, and Cameroon and is Africa’s most populous country, with over 230 million people, 54.9% of whom live in urban areas [35]. The country has a median age of 18.1 years, indicating a substantially young population [35]. Nigeria is highly diverse, with over 374 ethnic groups and languages. The three main groups—Hausa/Fulani, Yoruba, and Igbo, and several others—make the country one of the most multilingual nations in the world [36]. Administratively, Nigeria is divided into 36 states and the Federal Capital Territory (FCT), organised into six geopolitical zones (‘regions’): North-East (six states), North Central (six states and the FCT), North-West (seven states), South-South (six states), South-West (six states), and South-East (five states) [Fig 1]. Nigeria also has 774 local government areas (LGAs), with wards or enumeration areas serving as the smallest administrative units. Approximately 63% of Nigerians experience multidimensional poverty, reflecting deep socioeconomic inequalities [37].

**Fig 1:**
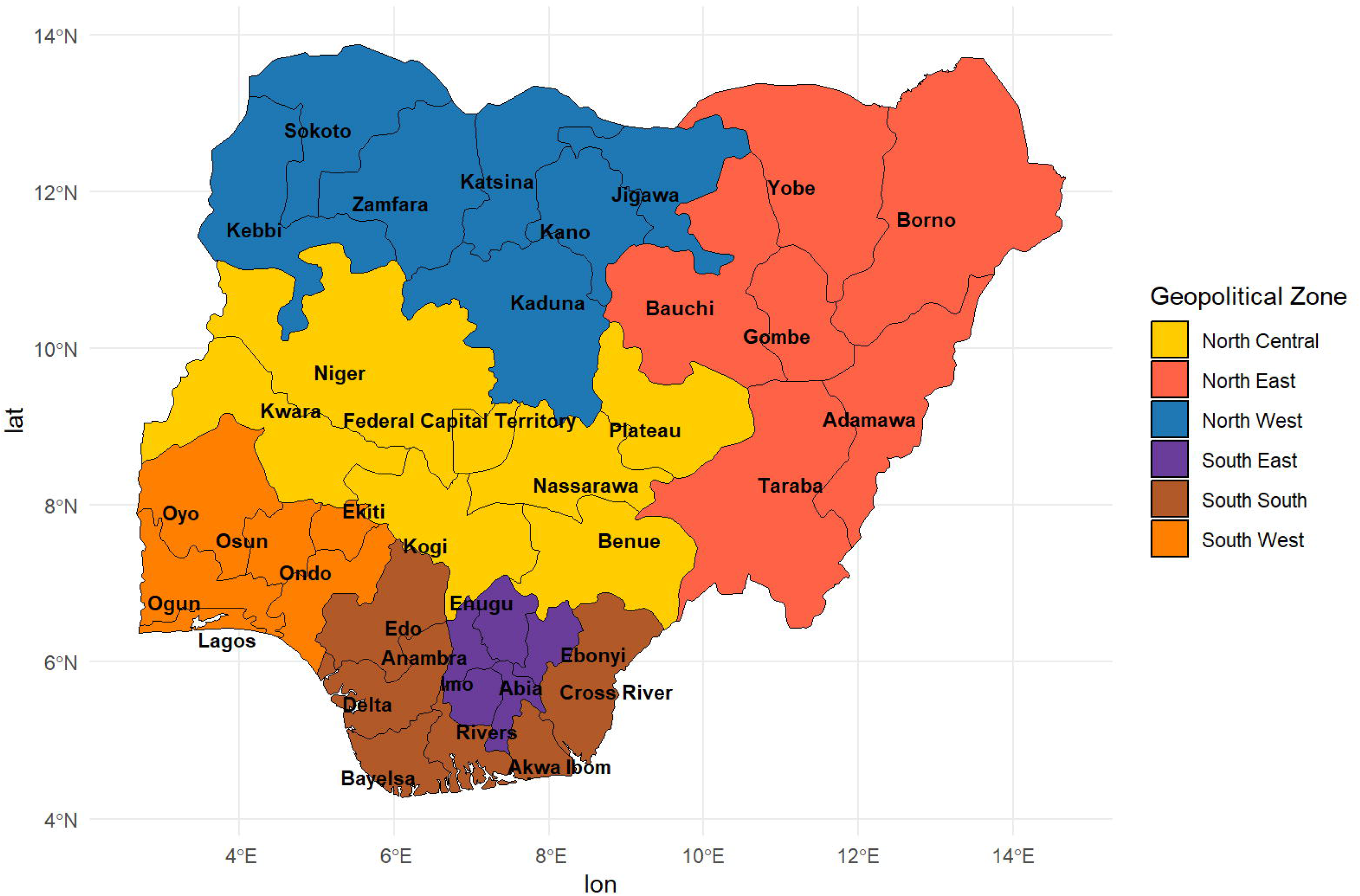
Map of Nigeria showing the geo-political zones.

Nigeria’s healthcare system comprises both public and private sectors, operating in parallel and with limited integration. The public sector follows a three-tier structure: primary healthcare managed at the local government level through primary health centres; secondary care provided by state-owned general hospitals; and tertiary care delivered through teaching hospitals and federal medical centres offering specialised services. Despite this structure, the private sector, including formal and informal providers, plays a major role in service delivery and accounts for a considerable proportion of healthcare utilisation in Nigeria [38]. Some initiatives aligned with global health goals have been implemented in Nigeria [8–11]; however, the country experiences poor maternal and neonatal health outcomes. Home birth is a potential contributor, and national data may obscure within-population disparities [24, 39, 40]; hence the approach in this study.

### Data Source and Sampling Design

We utilised data from the cross-sectional Nigerian Demographic and Health Survey (NDHS) 2018. The NDHS is a nationally representative survey conducted periodically since its inception in 1990 [13]. The survey employs a rigorously tested and validated methodology. The 2018 version was implemented by the Nigerian National Population Commission in collaboration with international partners and technical oversight from ICF International. The survey offers key demographic and health indicators, including fertility, family planning, maternal and child health, and health service utilisation. The NDHS 2018 used a stratified two-stage cluster sampling method to select 42,000 households from 1,400 clusters [13]. Data collection was through standardised, validated questionnaires administered by trained interviewers [13]. The completed survey covered 40,427 households within 1,389 clusters. A total of 41,821 mothers aged 15–49 participated (16,984 from urban and 24,837 from rural areas). The overall response rate was high at 99.3%, with urban and rural response rates of 99.2% and 99.4%, respectively.

This study analysed a weighted representative sample of 21,512 mothers—8,467 from urban areas and 13,055 from rural areas—who had a live birth in the five years before the survey [13]. These mothers provided complete data on the place of childbirth for their most recent live birth. We used the Children Recode (KR) dataset, restricted to the most recent live birth per mother. This dataset includes detailed records on pregnancy, delivery, postnatal care, and child health, alongside maternal data, fully anonymised to protect personally identifiable information. Documentation on the NDHS 2018 methodology is publicly available [13]. Ethical approval was granted by the Nigerian National Health Research Ethics Committee, and informed consent was obtained from participants aged 18 and older, while parental consent was required for those under 18. The dataset can be accessed (https://dhsprogram.com/data/available-datasets.cfm) upon approval from the DHS Program [13].

### Study factors Outcome variable

The primary outcome variable was ‘home birth’ or ‘home delivery’, representing non-utilisation of a health facility or institution for childbirth. This outcome was derived from the ‘place of delivery’ variable, which was categorised into two groups: ‘home birth’ and ‘health facility birth.’ Based on our data, the ‘home birth’ category follows the practice in previous studies [19, 41], comprising births occurring at the ‘respondent’s home’ and ‘another home’. Conversely, health facility births encompassed childbirths occurring in public and private healthcare facilities. Public facilities included government hospitals, health centres, health posts, and other public sector institutions, while private facilities comprised private hospitals, clinics, and other privately operated healthcare providers/facilities.

### Explanatory variables

We adopt Andersen’s behavioural model as the conceptual framework and utilise the model in selecting explanatory variables—a well-established approach in healthcare services utilisation research [19, 24, 25, 42]. The model categorises factors influencing healthcare services use into three key domains: predisposing factors (demographic and social characteristics that influence healthcare-seeking behaviour), enabling factors (resources that facilitate or hinder access to healthcare), and need factors (individual perceptions of health needs) [31]. Using this framework, we examined how various factors are associated with having a birth at home rather than in a healthcare facility. The selection of explanatory variables was also guided by a review of previous research and the availability of data in the dataset [19, 24, 25, 41, 42]. These variables were grouped into four broad categories (Fig 2):

i. External environmental factors included respondents’ place of residence (urban or rural) and regions, with northern regions (North-West, North-Central and North-East) and southern regions (South-South, South-West, and South-East) classified separately (Figs 1 and 2).
ii. Predisposing factors were divided into health knowledge indicators (frequency of exposure to newspapers, television, radio, and the internet) and socio-demographic characteristics, including maternal age, household wealth, religion, education level, working status of the mother and her partner, decision-making autonomy, ethnicity, birth order, preceding birth interval, and birth type (Fig 2).
iii. Enabling factors encompass key aspects influencing healthcare access, such as health insurance, financial capacity to pay for services, distance to facility, and difficulties in obtaining permission to seek care.
iv. Need factors focused on pregnancy intention or desire for pregnancy, representing the mother’s intention at the time of conception.

Further details on the classification and operationalisation of these variables are in Table 1.

**Fig 2:**
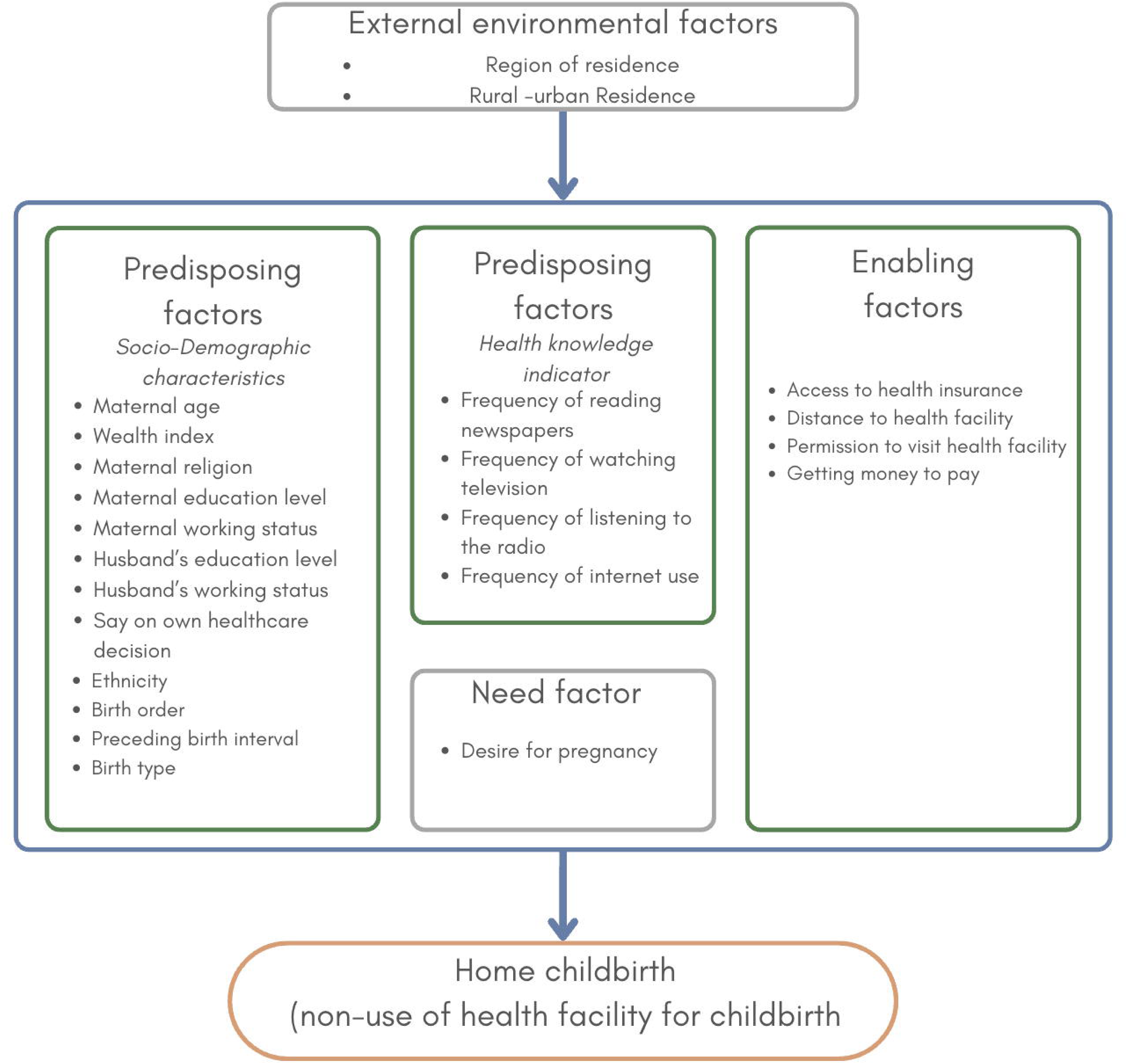
Theoretical framework for home birth in Nigeria (adapted from Andersen’s behavioural model)

**Table 1:**
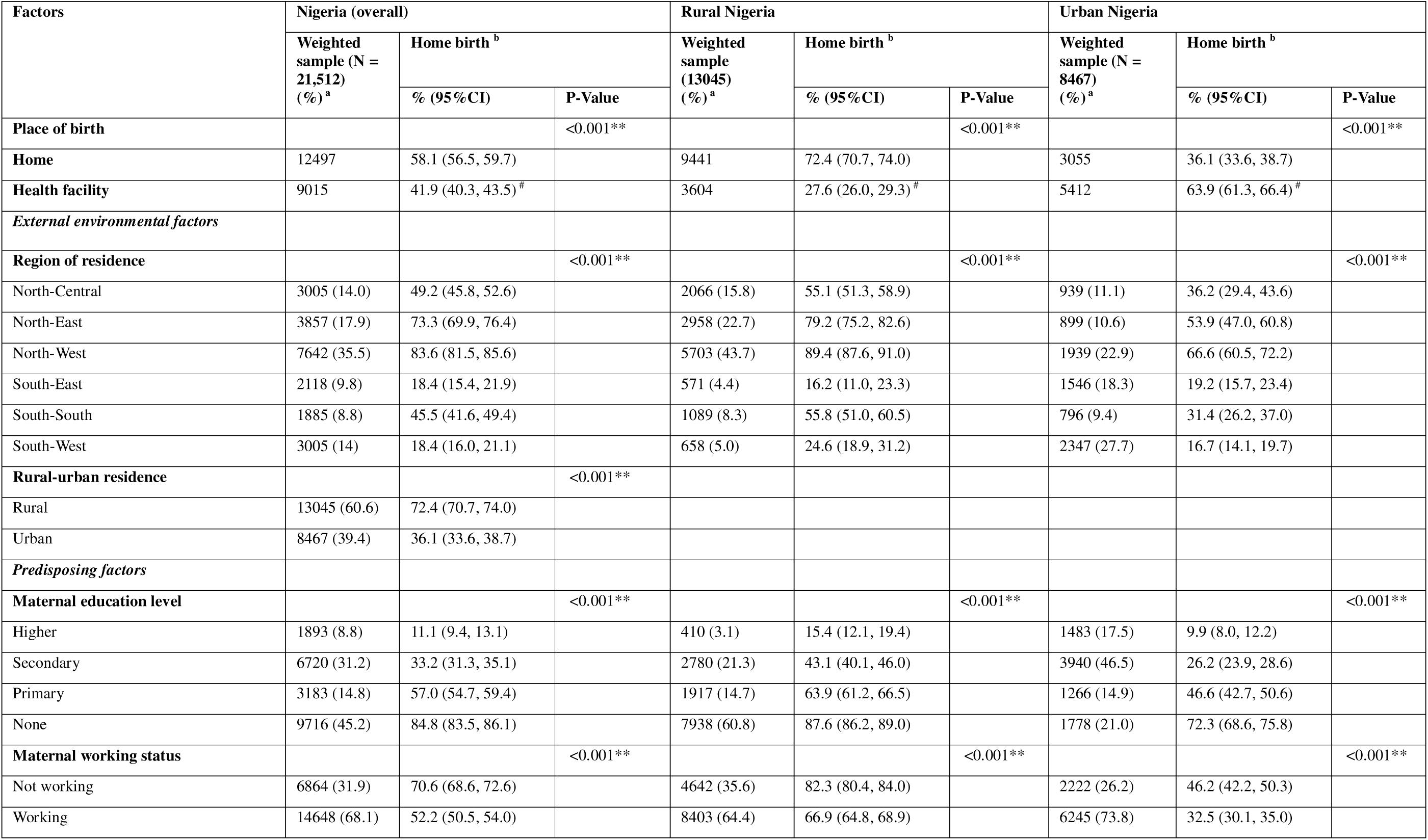
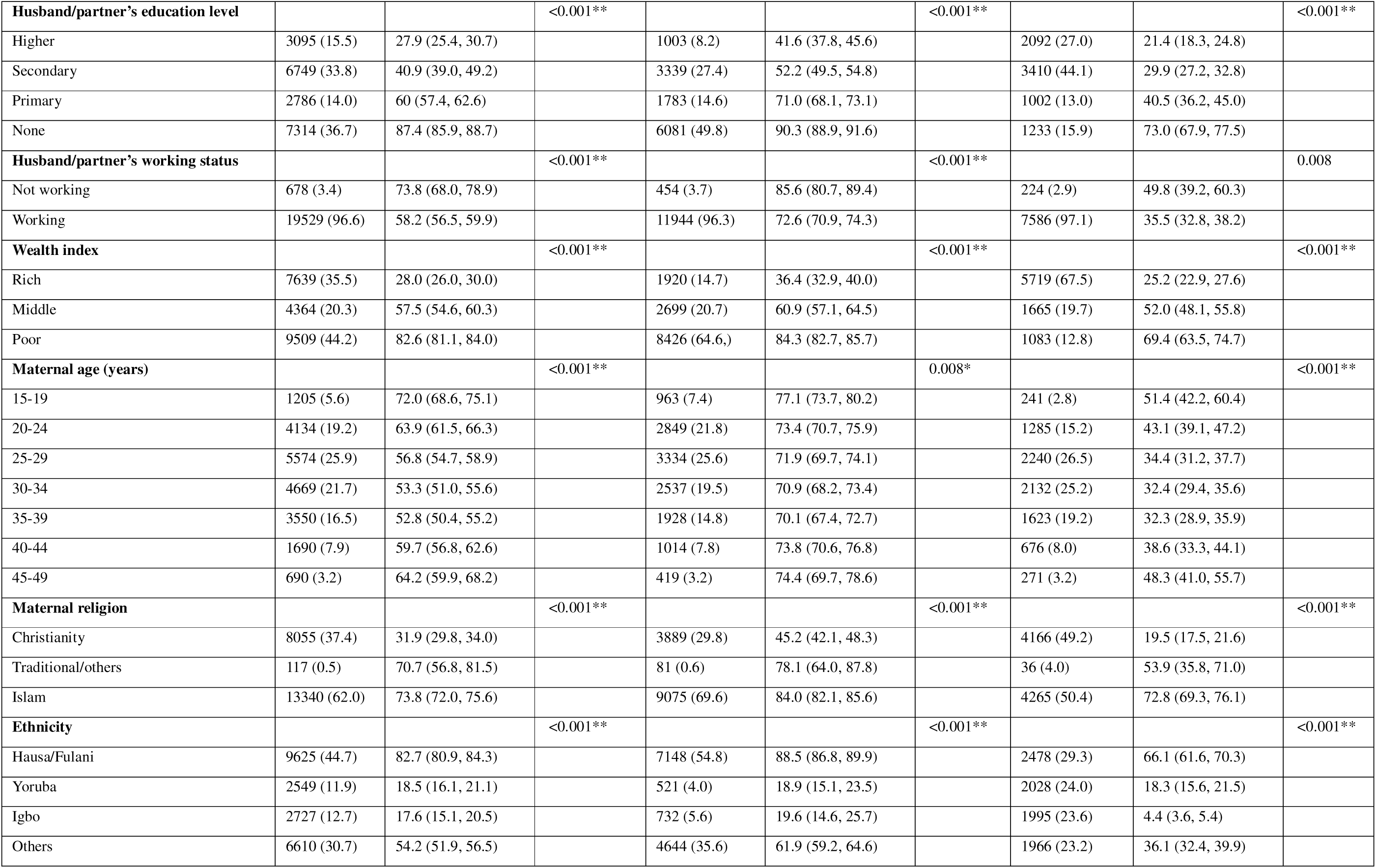
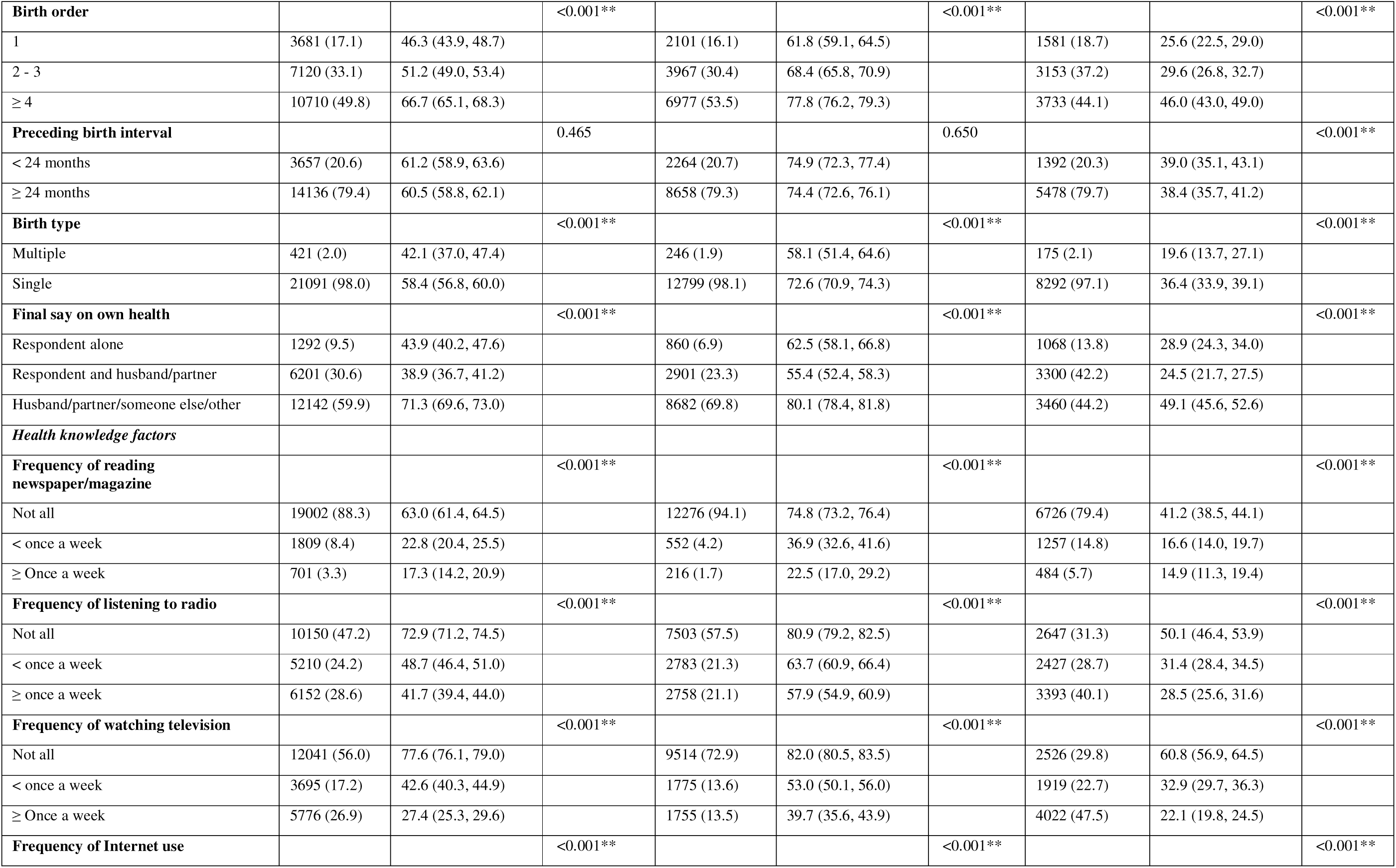
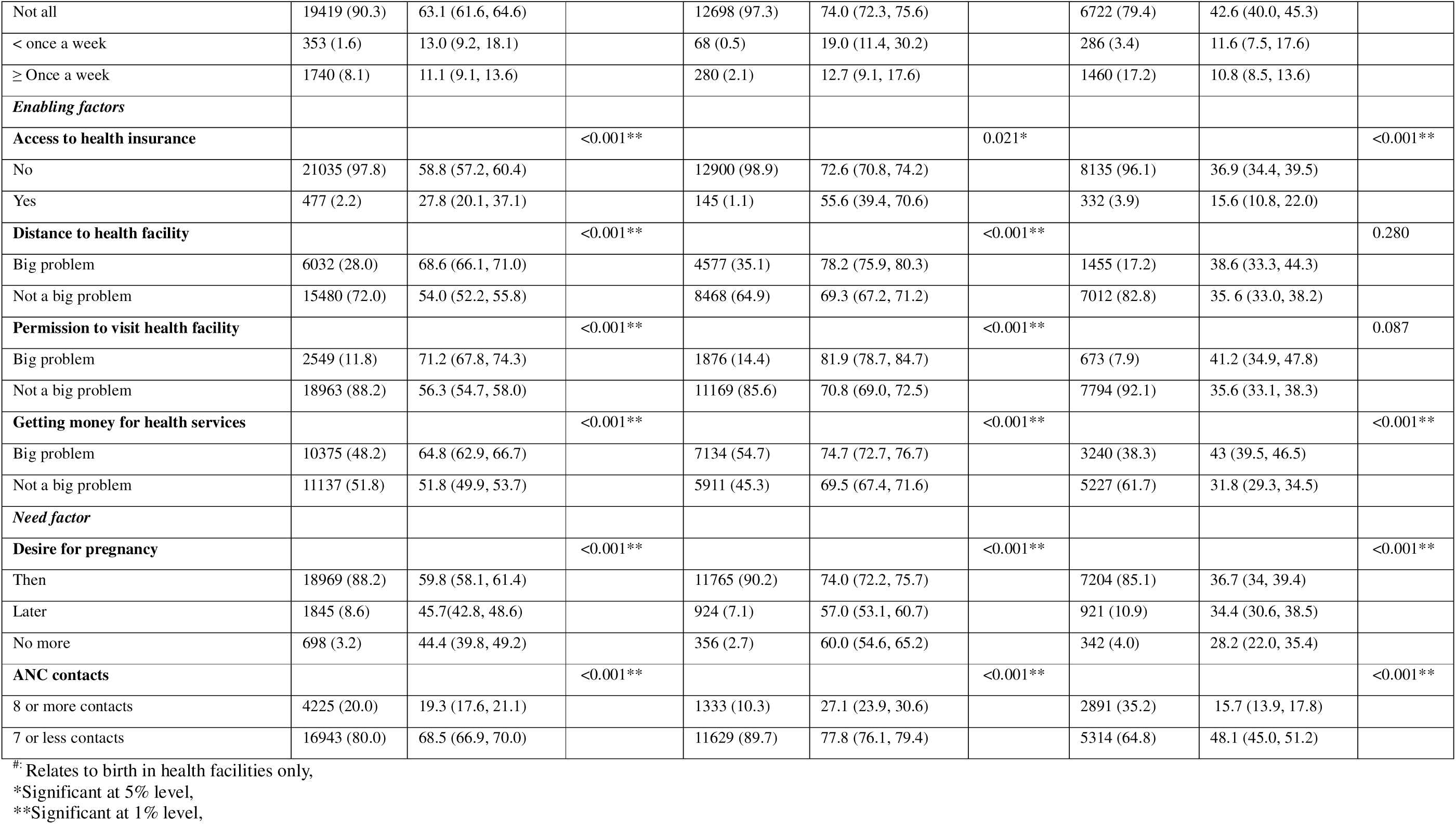
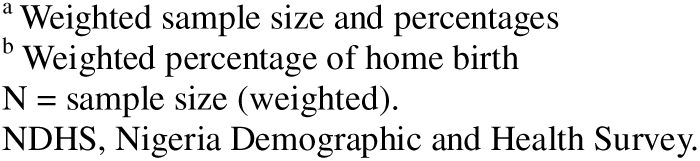
Sample characteristics and home birth in Nigeria by rural and urban residences, NDHS 2018.

| Factors | Nigeria (overall) |  |  | Rural Nigeria |  |  | Urban Nigeria |  |  |
| --- | --- | --- | --- | --- | --- | --- | --- | --- | --- |
|  | Weighted sample (N = 21,512) (%)<sup>a</sup> | Home birth<sup>b</sup> |  | Weighted sample (13045) (%)<sup>a</sup> | Home birth<sup>b</sup> |  | Weighted sample (N = 8467) (%)<sup>a</sup> | Home birth<sup>b</sup> |  |
|  |  | % (95%CI) | P-Value |  | % (95%CI) | P-Value |  | % (95%CI) | P-Value |
| **Place of birth** |  |  | <0.001** |  |  | <0.001** |  |  | <0.001** |
| **Home** | 12497 | 58.1 (56.5, 59.7) |  | 9441 | 72.4 (70.7, 74.0) |  | 3055 | 36.1 (33.6, 38.7) |  |
| **Health facility** | 9015 | 41.9 (40.3, 43.5)<sup>#</sup> |  | 3604 | 27.6 (26.0, 29.3)<sup>#</sup> |  | 5412 | 63.9 (61.3, 66.4)<sup>#</sup> |  |
| *External environmental factors* |  |  |  |  |  |  |  |  |  |
| **Region of residence** |  |  | <0.001** |  |  | <0.001** |  |  | <0.001** |
| North-Central | 3005 (14.0) | 49.2 (45.8, 52.6) |  | 2066 (15.8) | 55.1 (51.3, 58.9) |  | 939 (11.1) | 36.2 (29.4, 43.6) |  |
| North-East | 3857 (17.9) | 73.3 (69.9, 76.4) |  | 2958 (22.7) | 79.2 (75.2, 82.6) |  | 899 (10.6) | 53.9 (47.0, 60.8) |  |
| North-West | 7642 (35.5) | 83.6 (81.5, 85.6) |  | 5703 (43.7) | 89.4 (87.6, 91.0) |  | 1939 (22.9) | 66.6 (60.5, 72.2) |  |
| South-East | 2118 (9.8) | 18.4 (15.4, 21.9) |  | 571 (4.4) | 16.2 (11.0, 23.3) |  | 1546 (18.3) | 19.2 (15.7, 23.4) |  |
| South-South | 1885 (8.8) | 45.5 (41.6, 49.4) |  | 1089 (8.3) | 55.8 (51.0, 60.5) |  | 796 (9.4) | 31.4 (26.2, 37.0) |  |
| South-West | 3005 (14) | 18.4 (16.0, 21.1) |  | 658 (5.0) | 24.6 (18.9, 31.2) |  | 2347 (27.7) | 16.7 (14.1, 19.7) |  |
| **Rural-urban residence** |  |  | <0.001** |  |  |  |  |  |  |
| Rural | 13045 (60.6) | 72.4 (70.7, 74.0) |  |  |  |  |  |  |  |
| Urban | 8467 (39.4) | 36.1 (33.6, 38.7) |  |  |  |  |  |  |  |
| *Predisposing factors* |  |  |  |  |  |  |  |  |  |
| **Maternal education level** |  |  | <0.001** |  |  | <0.001** |  |  | <0.001** |
| Higher | 1893 (8.8) | 11.1 (9.4, 13.1) |  | 410 (3.1) | 15.4 (12.1, 19.4) |  | 1483 (17.5) | 9.9 (8.0, 12.2) |  |
| Secondary | 6720 (31.2) | 33.2 (31.3, 35.1) |  | 2780 (21.3) | 43.1 (40.1, 46.0) |  | 3940 (46.5) | 26.2 (23.9, 28.6) |  |
| Primary | 3183 (14.8) | 57.0 (54.7, 59.4) |  | 1917 (14.7) | 63.9 (61.2, 66.5) |  | 1266 (14.9) | 46.6 (42.7, 50.6) |  |
| None | 9716 (45.2) | 84.8 (83.5, 86.1) |  | 7938 (60.8) | 87.6 (86.2, 89.0) |  | 1778 (21.0) | 72.3 (68.6, 75.8) |  |
| **Maternal working status** |  |  | <0.001** |  |  | <0.001** |  |  | <0.001** |
| Not working | 6864 (31.9) | 70.6 (68.6, 72.6) |  | 4642 (35.6) | 82.3 (80.4, 84.0) |  | 2222 (26.2) | 46.2 (42.2, 50.3) |  |
| Working | 14648 (68.1) | 52.2 (50.5, 54.0) |  | 8403 (64.4) | 66.9 (64.8, 68.9) |  | 6245 (73.8) | 32.5 (30.1, 35.0) |  |
| **Husband/partner's education level** |  |  | <0.001** |  |  | <0.001** |  |  | <0.001** |
| Higher | 3095 (15.5) | 27.9 (25.4, 30.7) |  | 1003 (8.2) | 41.6 (37.8, 45.6) |  | 2092 (27.0) | 21.4 (18.3, 24.8) |  |
| Secondary | 6749 (33.8) | 40.9 (39.0, 49.2) |  | 3339 (27.4) | 52.2 (49.5, 54.8) |  | 3410 (44.1) | 29.9 (27.2, 32.8) |  |
| Primary | 2786 (14.0) | 60 (57.4, 62.6) |  | 1783 (14.6) | 71.0 (68.1, 73.1) |  | 1002 (13.0) | 40.5 (36.2, 45.0) |  |
| None | 7314 (36.7) | 87.4 (85.9, 88.7) |  | 6081 (49.8) | 90.3 (88.9, 91.6) |  | 1233 (15.9) | 73.0 (67.9, 77.5) |  |
| **Husband/partner's working status** |  |  | <0.001** |  |  | <0.001** |  |  | 0.008 |
| Not working | 678 (3.4) | 73.8 (68.0, 78.9) |  | 454 (3.7) | 85.6 (80.7, 89.4) |  | 224 (2.9) | 49.8 (39.2, 60.3) |  |
| Working | 19529 (96.6) | 58.2 (56.5, 59.9) |  | 11944 (96.3) | 72.6 (70.9, 74.3) |  | 7586 (97.1) | 35.5 (32.8, 38.2) |  |
| **Wealth index** |  |  | <0.001** |  |  | <0.001** |  |  | <0.001** |
| Rich | 7639 (35.5) | 28.0 (26.0, 30.0) |  | 1920 (14.7) | 36.4 (32.9, 40.0) |  | 5719 (67.5) | 25.2 (22.9, 27.6) |  |
| Middle | 4364 (20.3) | 57.5 (54.6, 60.3) |  | 2699 (20.7) | 60.9 (57.1, 64.5) |  | 1665 (19.7) | 52.0 (48.1, 55.8) |  |
| Poor | 9509 (44.2) | 82.6 (81.1, 84.0) |  | 8426 (64.6) | 84.3 (82.7, 85.7) |  | 1083 (12.8) | 69.4 (63.5, 74.7) |  |
| **Maternal age (years)** |  |  | <0.001** |  |  | 0.008* |  |  | <0.001** |
| 15-19 | 1205 (5.6) | 72.0 (68.6, 75.1) |  | 963 (7.4) | 77.1 (73.7, 80.2) |  | 241 (2.8) | 51.4 (42.2, 60.4) |  |
| 20-24 | 4134 (19.2) | 63.9 (61.5, 66.3) |  | 2849 (21.8) | 73.4 (70.7, 75.9) |  | 1285 (15.2) | 43.1 (39.1, 47.2) |  |
| 25-29 | 5574 (25.9) | 56.8 (54.7, 58.9) |  | 3334 (25.6) | 71.9 (69.7, 74.1) |  | 2240 (26.5) | 34.4 (31.2, 37.7) |  |
| 30-34 | 4669 (21.7) | 53.3 (51.0, 55.6) |  | 2537 (19.5) | 70.9 (68.2, 73.4) |  | 2132 (25.2) | 32.4 (29.4, 35.6) |  |
| 35-39 | 3550 (16.5) | 52.8 (50.4, 55.2) |  | 1928 (14.8) | 70.1 (67.4, 72.7) |  | 1623 (19.2) | 32.3 (28.9, 35.9) |  |
| 40-44 | 1690 (7.9) | 59.7 (56.8, 62.6) |  | 1014 (7.8) | 73.8 (70.6, 76.8) |  | 676 (8.0) | 38.6 (33.3, 44.1) |  |
| 45-49 | 690 (3.2) | 64.2 (59.9, 68.2) |  | 419 (3.2) | 74.4 (69.7, 78.6) |  | 271 (3.2) | 48.3 (41.0, 55.7) |  |
| **Maternal religion** |  |  | <0.001** |  |  | <0.001** |  |  | <0.001** |
| Christianity | 8055 (37.4) | 31.9 (29.8, 34.0) |  | 3889 (29.8) | 45.2 (42.1, 48.3) |  | 4166 (49.2) | 19.5 (17.5, 21.6) |  |
| Traditional/others | 117 (0.5) | 70.7 (56.8, 81.5) |  | 81 (0.6) | 78.1 (64.0, 87.8) |  | 36 (4.0) | 53.9 (35.8, 71.0) |  |
| Islam | 13340 (62.0) | 73.8 (72.0, 75.6) |  | 9075 (69.6) | 84.0 (82.1, 85.6) |  | 4265 (50.4) | 72.8 (69.3, 76.1) |  |
| **Ethnicity** |  |  | <0.001** |  |  | <0.001** |  |  | <0.001** |
| Hausa/Fulani | 9625 (44.7) | 82.7 (80.9, 84.3) |  | 7148 (54.8) | 88.5 (86.8, 89.9) |  | 2478 (29.3) | 66.1 (61.6, 70.3) |  |
| Yoruba | 2549 (11.9) | 18.5 (16.1, 21.1) |  | 521 (4.0) | 18.9 (15.1, 23.5) |  | 2028 (24.0) | 18.3 (15.6, 21.5) |  |
| Igbo | 2727 (12.7) | 17.6 (15.1, 20.5) |  | 732 (5.6) | 19.6 (14.6, 25.7) |  | 1995 (23.6) | 4.4 (3.6, 5.4) |  |
| Others | 6610 (30.7) | 54.2 (51.9, 56.5) |  | 4644 (35.6) | 61.9 (59.2, 64.6) |  | 1966 (23.2) | 36.1 (32.4, 39.9) |  |
| **Birth order** |  |  | <0.001** |  |  | <0.001** |  |  | <0.001** |
| 1 | 3681 (17.1) | 46.3 (43.9, 48.7) |  | 2101 (16.1) | 61.8 (59.1, 64.5) |  | 1581 (18.7) | 25.6 (22.5, 29.0) |  |
| 2 - 3 | 7120 (33.1) | 51.2 (49.0, 53.4) |  | 3967 (30.4) | 68.4 (65.8, 70.9) |  | 3153 (37.2) | 29.6 (26.8, 32.7) |  |
| ≥ 4 | 10710 (49.8) | 66.7 (65.1, 68.3) |  | 6977 (53.5) | 77.8 (76.2, 79.3) |  | 3733 (44.1) | 46.0 (43.0, 49.0) |  |
| **Preceding birth interval** |  |  | 0.465 |  |  | 0.650 |  |  | <0.001** |
| < 24 months | 3657 (20.6) | 61.2 (58.9, 63.6) |  | 2264 (20.7) | 74.9 (72.3, 77.4) |  | 1392 (20.3) | 39.0 (35.1, 43.1) |  |
| ≥ 24 months | 14136 (79.4) | 60.5 (58.8, 62.1) |  | 8658 (79.3) | 74.4 (72.6, 76.1) |  | 5478 (79.7) | 38.4 (35.7, 41.2) |  |
| **Birth type** |  |  | <0.001** |  |  | <0.001** |  |  | <0.001** |
| Multiple | 421 (2.0) | 42.1 (37.0, 47.4) |  | 246 (1.9) | 58.1 (51.4, 64.6) |  | 175 (2.1) | 19.6 (13.7, 27.1) |  |
| Single | 21091 (98.0) | 58.4 (56.8, 60.0) |  | 12799 (98.1) | 72.6 (70.9, 74.3) |  | 8292 (97.1) | 36.4 (33.9, 39.1) |  |
| **Final say on own health** |  |  | <0.001** |  |  | <0.001** |  |  | <0.001** |
| Respondent alone | 1292 (9.5) | 43.9 (40.2, 47.6) |  | 860 (6.9) | 62.5 (58.1, 66.8) |  | 1068 (13.8) | 28.9 (24.3, 34.0) |  |
| Respondent and husband/partner | 6201 (30.6) | 38.9 (36.7, 41.2) |  | 2901 (23.3) | 55.4 (52.4, 58.3) |  | 3300 (42.2) | 24.5 (21.7, 27.5) |  |
| Husband/partner/someone else/other | 12142 (59.9) | 71.3 (69.6, 73.0) |  | 8682 (69.8) | 80.1 (78.4, 81.8) |  | 3460 (44.2) | 49.1 (45.6, 52.6) |  |
| **Health knowledge factors** |  |  |  |  |  |  |  |  |  |
| **Frequency of reading newspaper/magazine** |  |  | <0.001** |  |  | <0.001** |  |  | <0.001** |
| Not all | 19002 (88.3) | 63.0 (61.4, 64.5) |  | 12276 (94.1) | 74.8 (73.2, 76.4) |  | 6726 (79.4) | 41.2 (38.5, 44.1) |  |
| < once a week | 1809 (8.4) | 22.8 (20.4, 25.5) |  | 552 (4.2) | 36.9 (32.6, 41.6) |  | 1257 (14.8) | 16.6 (14.0, 19.7) |  |
| ≥ Once a week | 701 (3.3) | 17.3 (14.2, 20.9) |  | 216 (1.7) | 22.5 (17.0, 29.2) |  | 484 (5.7) | 14.9 (11.3, 19.4) |  |
| **Frequency of listening to radio** |  |  | <0.001** |  |  | <0.001** |  |  | <0.001** |
| Not all | 10150 (47.2) | 72.9 (71.2, 74.5) |  | 7503 (57.5) | 80.9 (79.2, 82.5) |  | 2647 (31.3) | 50.1 (46.4, 53.9) |  |
| < once a week | 5210 (24.2) | 48.7 (46.4, 51.0) |  | 2783 (21.3) | 63.7 (60.9, 66.4) |  | 2427 (28.7) | 31.4 (28.4, 34.5) |  |
| ≥ once a week | 6152 (28.6) | 41.7 (39.4, 44.0) |  | 2758 (21.1) | 57.9 (54.9, 60.9) |  | 3393 (40.1) | 28.5 (25.6, 31.6) |  |
| **Frequency of watching television** |  |  | <0.001** |  |  | <0.001** |  |  | <0.001** |
| Not all | 12041 (56.0) | 77.6 (76.1, 79.0) |  | 9514 (72.9) | 82.0 (80.5, 83.5) |  | 2526 (29.8) | 60.8 (56.9, 64.5) |  |
| < once a week | 3695 (17.2) | 42.6 (40.3, 44.9) |  | 1775 (13.6) | 53.0 (50.1, 56.0) |  | 1919 (22.7) | 32.9 (29.7, 36.3) |  |
| ≥ Once a week | 5776 (26.9) | 27.4 (25.3, 29.6) |  | 1755 (13.5) | 39.7 (35.6, 43.9) |  | 4022 (47.5) | 22.1 (19.8, 24.5) |  |
| **Frequency of Internet use** |  |  | <0.001** |  |  | <0.001** |  |  | <0.001** |
| Not all | 19419 (90.3) | 63.1 (61.6, 64.6) |  | 12698 (97.3) | 74.0 (72.3, 75.6) |  | 6722 (79.4) | 42.6 (40.0, 45.3) |  |
| < once a week | 353 (1.6) | 13.0 (9.2, 18.1) |  | 68 (0.5) | 19.0 (11.4, 30.2) |  | 286 (3.4) | 11.6 (7.5, 17.6) |  |
| ≥ Once a week | 1740 (8.1) | 11.1 (9.1, 13.6) |  | 280 (2.1) | 12.7 (9.1, 17.6) |  | 1460 (17.2) | 10.8 (8.5, 13.6) |  |
| **Enabling factors** |  |  |  |  |  |  |  |  |  |
| **Access to health insurance** |  |  | <0.001** |  |  | 0.021* |  |  | <0.001** |
| No | 21035 (97.8) | 58.8 (57.2, 60.4) |  | 12900 (98.9) | 72.6 (70.8, 74.2) |  | 8135 (96.1) | 36.9 (34.4, 39.5) |  |
| Yes | 477 (2.2) | 27.8 (20.1, 37.1) |  | 145 (1.1) | 55.6 (39.4, 70.6) |  | 332 (3.9) | 15.6 (10.8, 22.0) |  |
| **Distance to health facility** |  |  | <0.001** |  |  | <0.001** |  |  | 0.280 |
| Big problem | 6032 (28.0) | 68.6 (66.1, 71.0) |  | 4577 (35.1) | 78.2 (75.9, 80.3) |  | 1455 (17.2) | 38.6 (33.3, 44.3) |  |
| Not a big problem | 15480 (72.0) | 54.0 (52.2, 55.8) |  | 8468 (64.9) | 69.3 (67.2, 71.2) |  | 7012 (82.8) | 35.6 (33.0, 38.2) |  |
| **Permission to visit health facility** |  |  | <0.001** |  |  | <0.001** |  |  | 0.087 |
| Big problem | 2549 (11.8) | 71.2 (67.8, 74.3) |  | 1876 (14.4) | 81.9 (78.7, 84.7) |  | 673 (7.9) | 41.2 (34.9, 47.8) |  |
| Not a big problem | 18963 (88.2) | 56.3 (54.7, 58.0) |  | 11169 (85.6) | 70.8 (69.0, 72.5) |  | 7794 (92.1) | 35.6 (33.1, 38.3) |  |
| **Getting money for health services** |  |  | <0.001** |  |  | <0.001** |  |  | <0.001** |
| Big problem | 10375 (48.2) | 64.8 (62.9, 66.7) |  | 7134 (54.7) | 74.7 (72.7, 76.7) |  | 3240 (38.3) | 43 (39.5, 46.5) |  |
| Not a big problem | 11137 (51.8) | 51.8 (49.9, 53.7) |  | 5911 (45.3) | 69.5 (67.4, 71.6) |  | 5227 (61.7) | 31.8 (29.3, 34.5) |  |
| **Need factor** |  |  |  |  |  |  |  |  |  |
| **Desire for pregnancy** |  |  | <0.001** |  |  | <0.001** |  |  | <0.001** |
| Then | 18969 (88.2) | 59.8 (58.1, 61.4) |  | 11765 (90.2) | 74.0 (72.2, 75.7) |  | 7204 (85.1) | 36.7 (34, 39.4) |  |
| Later | 1845 (8.6) | 45.7 (42.8, 48.6) |  | 924 (7.1) | 57.0 (53.1, 60.7) |  | 921 (10.9) | 34.4 (30.6, 38.5) |  |
| No more | 698 (3.2) | 44.4 (39.8, 49.2) |  | 356 (2.7) | 60.0 (54.6, 65.2) |  | 342 (4.0) | 28.2 (22.0, 35.4) |  |
| **ANC contacts** |  |  | <0.001** |  |  | <0.001** |  |  | <0.001** |
| 8 or more contacts | 4225 (20.0) | 19.3 (17.6, 21.1) |  | 1333 (10.3) | 27.1 (23.9, 30.6) |  | 2891 (35.2) | 15.7 (13.9, 17.8) |  |
| 7 or less contacts | 16943 (80.0) | 68.5 (66.9, 70.0) |  | 11629 (89.7) | 77.8 (76.1, 79.4) |  | 5314 (64.8) | 48.1 (45.0, 51.2) |  |

#. Relates to birth in health facilities only,
\*Significant at 5% level,
\*\*Significant at 1% level,
<sup>a</sup> Weighted sample size and percentages <sup>b</sup> Weighted percentage of home birth N = sample size (weighted). NDHS, Nigeria Demographic and Health Survey.

### Statistical analysis

We assessed both unadjusted (univariable) and adjusted (multivariable) relationships between home birth and selected explanatory variables using a two-step approach. First, we performed analyses using aggregated data for the overall Nigerian population, and second, we disaggregated our data and conducted separate analyses for urban and rural settings. In each setting, we estimated the proportion of home births, along with the corresponding 95% confidence intervals (CI), using frequency tabulation. Unadjusted associations between home birth and each explanatory factor were evaluated using Chi-square tests, and only variables with statistically significant associations (p < 0.05) were advanced to the multivariable analyses.

For the multivariable analyses, we built logistic regression models for all four variable categories and employed a backward elimination strategy to systematically remove non-significant variables, retaining only those that were statistically significant at 5% level. Before fitting the models, we assessed multicollinearity among independent variables using Variance Inflation Factors (VIFs) and tolerance values, with VIFs <10 and tolerance >0.1 considered acceptable thresholds [43]. We further validated the robustness of the final parsimonious model by testing it against potential confounders and factors previously reported as being associated with home birth. All analyses excluded responses marked as ‘missing’ or ‘don’t know’. We conducted all statistical analyses and data management in IBM SPSS Statistics for Windows (Version 21.0), using its Complex Samples module to account for sampling weights, stratification, and clustering in the NDHS 2018 design, ensuring that our estimates accurately reflect the survey’s complex sampling structure.

## Results

### Sample characteristics for the overall Nigerian data

The overall weighted sample consisted of 21,512 mothers aged 15–49 years. Regionally, the North-West had the highest representation at 35.5%, while the South-South had the lowest at 8.8% [Table 1]. Nearly half of the mothers (45.2%) had no formal education, while 8.8% had higher education. In terms of age, the cumulative percentage reaches just over 50% in the 25–29 age group. Using the grouped data formula, the estimated median age was 29.9 years. Teenagers comprised 5.6% of the sample, while most mothers were between 20 and 34 years (66.8%), highlighting a predominantly young-middle-aged population. Additionally, 68.1% of the mothers were working, 35.5% were in rich households, while 44.2% were in poor households.

### Sample characteristics for rural and urban residences

Rural-urban data disaggregation revealed distinct differences (Table 1). Of the total sample, 60.6% (13,045) resided in rural areas and 39.4% (8,467) in urban areas. In rural areas, a higher proportion of mothers had no formal education (60.8%) compared to urban mothers (21.0%). Working status also differed, with 64.4% of rural mothers engaged in work-related activities compared to 73.8% of urban mothers. However, urban residents typically had higher educational attainment (17.5% with higher education) than their rural counterparts (3.21%), suggesting variations in the type and quality of work. In urban areas, 67.5% of mothers were in the rich category, and only 12.8% were in the poor category. In contrast, just 14.7% of rural mothers were rich, and 64.6% were in the poor category.

### Prevalence of home birth in Nigeria by rural and urban residence

Overall, 58.1% (95% CI: 56.5, 59.7) of mothers in Nigeria gave birth at home, while 41.9% (95% CI: 40.3, 43.5) delivered at a health facility (Fig 3). Home birth prevalence was twice as high in rural (72.4% [95% CI: 70.7, 74.0]) as in urban areas (36.1% [95% CI: 33.6, 38.7]). At the national level, the North-West had the highest home birth prevalence (83.6% [95% CI: 81.5, 85.6]), while the South-East (18.4%; 95% CI: 15.4, 21.9) and South-West (18.4%; 95% CI: 16.0, 21.1) had the lowest.

**Fig 3:**
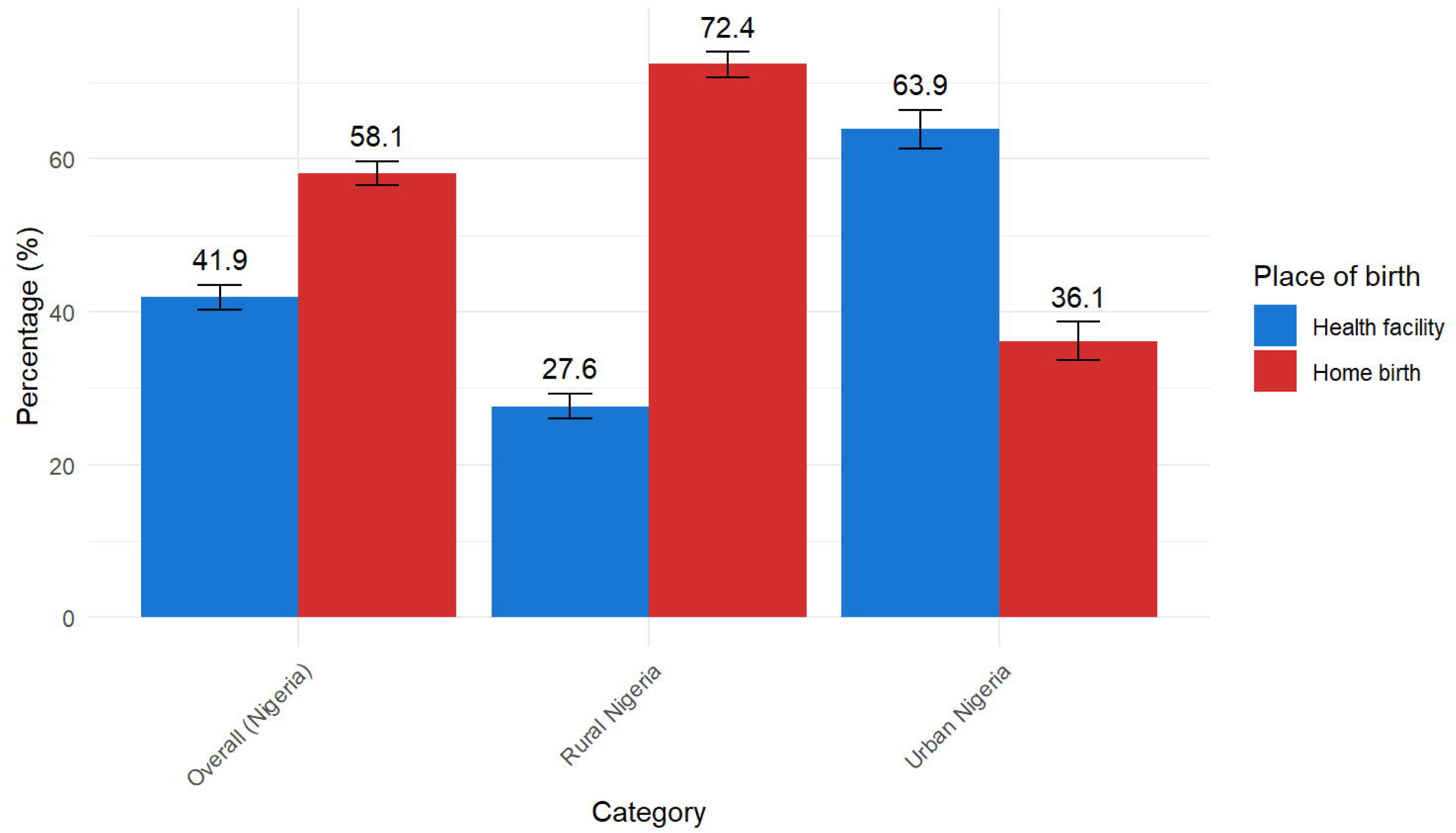
Home and health facility birth prevalence in Nigeria by national, rural and urban contexts.

Considering rural–urban residence-specific differences, the South-East had the lowest home birth prevalence in rural areas (16.2%; 95% CI: 11.0, 23.3), while the South-West had the lowest in urban areas (16.7%; 95% CI: 14.1, 19.7) (Fig 4). Conversely, the North-West recorded the highest prevalence, with 89.4% (95% CI: 87.6, 91.0) in rural areas and 66.6% (95% CI: 60.5, 72.2) in urban areas. Across all regions—except the South-East—rural areas consistently reported a higher home birth prevalence than urban areas. Surprisingly, in the South-East region, home birth was slightly higher (but not statistically significant) in urban settings (19.2%; 95% CI: 15.7, 23.4) than in rural areas (16.2%; 95% CI: 11.0, 23.3). Overall, the southern regions exhibited lower home birth prevalence than the northern regions (Fig 4). In all settings, the Hausa/Fulani ethnic group had the highest home birth prevalence. Also, mothers who had no education, identified as Muslim, had fewer than eight antenatal care contacts, were teenagers, or belonged to the low wealth index category had higher home birth prevalence.

**Fig 4:**
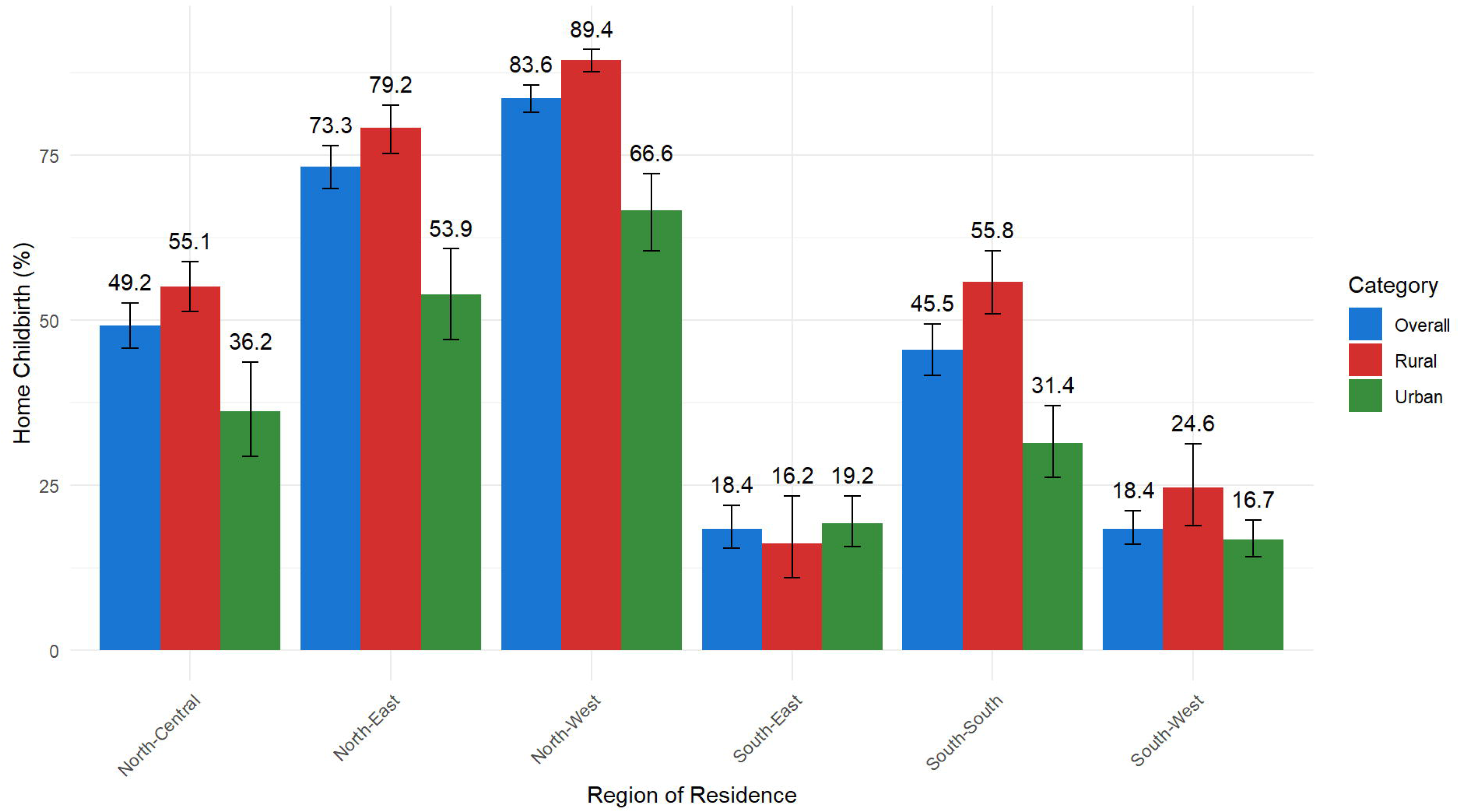
Home birth in Nigeria’s geopolitical zones by national, rural and urban residences.

### Factors associated with home birth in the overall Nigerian population

All VIFs were below five, and all tolerance values exceeded 0.1, indicating no significant multicollinearity among the predictor variables. Mothers residing in the North-East of Nigeria (AORlll=lll1.87; 95%CI: 1.34, 2.61), North-West (AORlll=lll3.30; 95%CI: 2.36, 4.64), and South-South (AORlll=lll4.54; 95%CI: 3.21, 6.43) had significantly higher odds of home birth compared to the South-West (Table 2). Mothers with no education (AORlll=lll3.18; 95% CI: 2.35, 4.30), primary education (AORlll=lll2.41; 95% CI: 1.80, 3.23), or secondary education (AORlll=lll1.87; 95% CI: 1.43, 2.45) had greater odds of home birth in comparison with higher education. A similar pattern of results was observed for the partner’s education level.

**Table 2:** Factors associated with home birth in Nigeria across the overall, rural and urban areas.

| Factors | Overall |  | Rural |  | Urban |  |
| --- | --- | --- | --- | --- | --- | --- |
|  | AOR | 95%CI | AOR | 95%CI | AOR | 95%CI |
| **Region** |  |  |  |  |  |  |
| North-Central | 1.28 | 0.93, 1.76 | 1.77 | 1.07, 2.94 | 1.07 | 0.72, 1.58 |
| North-East | 1.87 | 1.34, 2.61 | 3.11 | 1.87, 5.19 | 1.3 | 0.83, 2.02 |
| North-West | 3.3 | 2.36, 4.64 | 5.83 | 3.43, 9.90 | 2.09 | 1.38, 3.17 |
| South-East | 1.44 | 0.87, 2.40 | 1.63 | 0.75, 3.54 | 1.25 | 0.66, 2.38 |
| South-South | 4.54 | 3.21, 6.43 | 7.82 | 4.55, 13.44 | 2.71 | 1.74, 4.20 |
| South-West | 1 | Reference | 1 | Reference | 1 | Reference |
| **Maternal education level** |  |  |  |  |  |  |
| No education | 3.18 | 2.35, 4.30 | 3.46 | 2.33, 5.12 | 3.1 | 2.03, 4.75 |
| Primary | 2.41 | 1.80, 3.23 | 2.47 | 1.69, 3.60 | 2.67 | 1.78, 4.01 |
| Secondary | 1.87 | 1.43, 2.45 | 1.97 | 1.38, 2.79 | 1.88 | 1.30, 2.70 |
| Higher | 1 | Reference | 1 | Reference | 1 | Reference |
| **Frequency of watching TV** |  |  |  |  |  |  |
| Not at all | 1.42 | 1.22, 1.64 | 1.26 | 1.00, 1.59 | 1.56 | 1.28, 1.89 |
| < once a week | 1.1 | 0.94, 1.28 | 0.93 | 0.74, 1.16 | 1.23 | 1.00, 1.51 |
| ≥ once a week | 1 | Reference | 1 | Reference | 1 | Reference |
| **Frequency of listening to radio** |  |  |  |  |  |  |
| Not at all | 1.36 | 1.18, 1.56 | 1.35 | 1.14, 1.60 | 1.3 | 1.04, 1.62 |
| < once a week | 1.14 | 1.00, 1.31 | 1.23 | 1.02, 1.49 | 1.08 | 0.89, 1.31 |
| ≥ once a week | 1 | Reference | 1 | Reference | 1 | Reference |
| **Husband education level** |  |  |  |  |  |  |
| No Education | 2.26 | 1.84, 2.77 | 2.77 | 2.18, 3.53 | 1.57 | 1.11, 2.21 |
| Primary | 1.55 | 1.29, 1.86 | 1.76 | 1.39, 2.21 | 1.34 | 1.01, 1.77 |
| Secondary | 1.3 | 1.11, 1.52 | 1.34 | 1.10, 1.63 | 1.24 | 0.98, 1.57 |
| Higher | 1 | Reference | 1 | Reference | 1 | Reference |
| **Permission to visit health facility** |  |  |  |  |  |  |
| Big problem | 1.22 | 1.03, 1.44 | 1.45 | 1.15, 1.82 |  |  |
| Not a big problem | 1 | Reference | 1 | Reference |  |  |
| **Distance to health facility** |  |  |  |  |  |  |
| Big problem | 1.29 | 1.14, 1.46 | 1.37 | 1.18, 1.60 |  |  |
| Not a big problem | 1 | Reference | 1 | Reference |  |  |
| **Maternal age** |  |  |  |  |  |  |
| 15-19 | 1.49 | 1.09, 2.05 |  |  | 2.19 | 1.25, 3.82 |
| 20-24 | 1.50 | 1.17, 1.92 |  |  | 1.79 | 1.21, 2.64 |
| 25-29 | 1.43 | 1.14, 1.79 |  |  | 1.65 | 1.15, 2.36 |
| 30-34 | 1.26 | 1.01, 1.58 |  |  | 1.52 | 1.06, 2.16 |
| 35-39 | 1.17 | 0.94, 1.47 |  |  | 1.32 | 0.92, 1.87 |
| 40-44 | 1.22 | 0.92, 1.61 |  |  | 1.42 | 0.91, 2.23 |
| 45-49 | 1 | Reference |  |  | 1 | Reference |
| **Wealth index** |  |  |  |  |  |  |
| Poor | 2.19 | 1.85, 2.59 | 1.8 | 1.43, 2.27 | 2.17 | 1.64, 2.87 |
| Middle | 1.35 | 1.17, 1.54 | 1.2 | 0.98, 1.46 | 1.46 | 1.22, 1.75 |
| Rich | 1 | Reference | 1 | Reference | 1 | Reference |
| **Birth type** |  |  |  |  |  |  |
| Single | 2.27 | 1.65, 3.12 | 1.94 | 1.29, 2.91 | 3.07 | 1.71, 5.51 |
| Multiple | 1 | Reference | 1 | Reference | 1 | Reference |
| **Frequency of using Internet** |  |  |  |  |  |  |
| Not at all | 1.51 | 1.12, 2.02 | 2.53 | 1.54, 4.15 | 2.41 | 1.33, 4.36 |
| < Once a week | 0.74 | 0.45, 1.22 | 1.76 | 0.68, 4.60 | 1.81 | 1.00, 3.28 |
| ≥ Once a week | 1 | Reference | 1 | Reference | 1 | Reference |
| **Birth order** |  |  |  |  |  |  |
| 2 - 3 | 1.51 | 1.28, 1.78 | 1.58 | 1.32, 1.89 | 1.42 | 1.07, 1.89 |
| ≥ 4 | 1.75 | 1.45, 2.12 | 1.65 | 1.41, 1.94 | 1.68 | 1.21, 2.34 |
| 1 | 1 | Reference | 1 | Reference | 1 | Reference |
| **Religion** |  |  |  |  |  |  |
| Traditionalist/others | 1.45 | 0.88, 2.40 |  |  | 2.05 | 0.97, 4.36 |
| Islam | 1.32 | 1.10, 1.58 |  |  | 1.55 | 1.19, 2.02 |
| Christianity | 1 | Reference |  |  | 1 | Reference |
| **Ethnicity** |  |  |  |  |  |  |
| Hausa/Fulani | 1.53 | 1.26, 1.87 | 1.52 | 1.16, 2.00 | 1.63 | 1.25, 2.13 |
| Yoruba | 1.02 | 0.72, 1.45 | 0.86 | 0.51, 1.46 | 0.99 | 0.65, 1.50 |
| Igbo | 0.51 | 0.35, 0.75 | 0.48 | 0.29, 0.79 | 0.54 | 0.33, 0.88 |
| Others | 1 | Reference | 1 | Reference | 1 | Reference |
| **Antenatal contacts** |  |  |  |  |  |  |
| < 7 ANC contacts (underuse) | 2.35 | 2.02, 2.74 | 3.03 | 2.43, 3.78 | 1.91 | 1.57, 2.32 |
| ≥8 ANC contacts (use) | 1 | Reference | 1 | Reference | 1 | Reference |
| **Say on own health** |  |  |  |  |  |  |
| Respondent alone |  |  | 0.73 | 0.59, 0.91 |  |  |
| Respondent and spouse |  |  | 0.83 | 0.72, 0.96 |  |  |
| Spouse alone/someone else/others |  |  | 1 | Reference |  |  |
| **Getting money for health services** |  |  |  |  |  |  |
| Big problem |  |  |  |  | 1.25 | 1.07, 1.47 |
| Not a big problem |  |  |  |  | 1 | Reference |

AOR: Adjusted odds ratio, ANC: Antenatal care CI: confidence interval

Mothers who did not watch television had higher odds of choosing home birth (AOR = 1.42; 95% CI: 1.22, 1.64) compared to those who watched at least once weekly. Similarly, those who did not listen to the radio at all had increased odds of home birth (AOR = 1.36; 95% CI: 1.18, 1.56). Mothers facing challenges in accessing health services also showed higher odds of home birth. Specifically, those who found obtaining permission to visit a facility problematic (AOR = 1.22; 95% CI: 1.03, 1.44) and those concerned about distance to a facility (AOR = 1.29; 95% CI: 1.14, 1.46) had increased odds of home birth. Additionally, mothers under 35 had higher odds of home birth compared to older age groups.

Notably, in a stepwise pattern, mothers from poor households (AORlll=lll2.19; 95% CI: 1.85–2.59) and those in the middle wealth category (AORlll=lll1.35; 95% CI: 1.17–1.54) had greater odds of a home birth compared to those from rich households. In addition, giving birth to a singleton, rather than multiples, was associated with increased odds of home birth. Not using the internet at all increased the odds of home birth (AORlll=lll1.51; 95% CI: 1.12–2.02). Higher birth order, compared to first births, and identifying as Muslim, compared to Christian, were also associated with elevated odds. Belonging to the Hausa/Fulani ethnic group increased the odds of home birth, while Igbo ethnicity was associated with substantially lower odds (Table 2). Finally, ANC contact was inversely related to home birth: mothers who had fewer than seven ANC visits had markedly higher odds compared to those with eight or more contacts.

### Similarities in factors associated with home birth between rural and urban settings

In both rural and urban settings, mothers with no education had significantly higher odds of home birth compared to those with higher education. The effect was slightly stronger in rural areas than in urban areas, but remained comparable (Table 2). Similarly, lower educational attainment at the primary and secondary levels was associated with increased odds of home birth in both settings. In both contexts, mothers from poor households also had greater odds of home birth compared to those from rich households (Table 2). A lack of partner’s education was associated with higher odds of home birth in both settings, with a slightly stronger effect in rural areas (AORlll=lll2.77; 95% CI: 2.18–3.53) than in urban areas (AORlll=lll1.57; 95% CI: 1.11–2.21). Mothers who recorded fewer than seven antenatal contacts had higher odds of home birth in both settings, with a stronger association in rural areas (AORlll=lll3.03; 95% CI: 2.43–3.78) than in urban areas (AORlll=lll1.91; 95% CI: 1.57–2.32). Additionally, higher birth order, Hausa-Fulani ethnicity, singleton births, and limited media exposure (TV, radio, and internet) were common factors associated with home birth across both rural and urban areas.

### Differences in factors associated with home birth between rural and urban settings

Notable differences emerged when comparing rural and urban residences. First, the odds of home birth were significantly higher in four rural regions: North-East, North-Central, North-West, and South-South, compared to the South-West, and by extension, the South-East. That is, in rural Nigeria, all the northern regions (North-Central, North-West and North-East), and only one southern region (South-South) had higher odds of home birth. Conversely, in urban areas, higher odds were observed in only two regions: one from the northern area (North-West) and one from the southern area (South-South). Regional disparities were more pronounced in rural areas. For example, the North-West region was associated with higher odds of home birth in both settings, but the effect was over twice as strong in rural (AORlll=lll5.83; 95% CI: 3.43–9.90) than urban areas (AORlll=lll2.09; 95% CI: 1.38–3.17). Similarly, in the South-South region, rural mothers had nearly three-fold higher odds (AORlll=lll7.82; 95% CI: 4.55–13.44) compared to urban counterparts (AORlll=lll2.71; 95% CI: 1.74–4.20).

Secondly, certain access-related factors were uniquely associated with rural areas. Mothers who had difficulty obtaining permission to visit a health facility had higher odds of home birth (AORlll=lll1.45; 95% CI: 1.15–1.82), as did those reporting distance as a major barrier (AORlll=lll1.37; 95% CI: 1.18– 1.60). Home birth odds were also higher among mothers not involved in healthcare decision-making. Specifically, those deciding alone or jointly with partners had lower odds of home birth, a pattern observed only in rural areas. Thirdly, in urban areas, maternal age, religion, and financial access were uniquely associated with home birth. Younger mothers, particularly those aged 15–19 (AORlll=lll2.19; 95% CI: 1.25–3.82) and 20–24 (AORlll=lll1.79; 95% CI: 1.21–2.65), had higher odds compared to older groups. Religion was significant only in urban settings; Muslim mothers had higher odds than Christians (AORlll=lll1.55; 95% CI: 1.19–2.02). Financial constraints also mattered in urban areas, where mothers who reported difficulty affording care had higher odds of home birth (AORlll=lll1.25; 95% CI: 1.07–1.47).

## Discussion

Our findings revealed a high national prevalence, with more than half of mothers (58%) giving birth at home in Nigeria. We observed marked differences by rural–urban residence, region, and socio-demographic characteristics. The rural–urban divide is particularly striking: nearly three-quarters (72%) of rural mothers gave birth at home, compared to just over one-third (36%) of urban mothers. These figures demonstrate the widespread nature of home births in Nigeria, with rural areas recording more than twice the prevalence estimated in urban settings. The national prevalence we observed (58%) is slightly lower than the 63% reported in 2013 [19, 23], but remains substantially higher than the estimates from several comparable African countries, such as Benin (14%)[44, 45], Ghana (21%)[46], and Mali (32.9%)[45], and twice as high as the global average of 28% [47]. Similarly, rural home birth prevalence declined modestly from 78% to 72%, and urban prevalence from 38% to 36% [19]. Despite these marginal improvements, the persistently high burden of home births in Nigeria is concerning. The observed reductions in our study fall short of global targets advocating for skilled attendance at all births, with the proportion of deliveries attended by skilled personnel serving as a critical indicator for achieving SDG 3.1 [1, 2]. The continued high prevalence is notable, as home births typically occur in Nigeria without emergency obstetric care or, in some cases, without anyone in attendance [48, 49].

We found wide regional disparities, with prevalence ranging from 84% in the North-West to 18% in the South-East and South-West regions. Even among urban areas, two-thirds (67%) of mothers in the urban North-West region gave birth at home, indicating that urban residence alone does not guarantee access to, or use of, health facilities for childbirth. These regional patterns reinforce the high national burden and the substantial within-country disparities. The persistently high prevalence in vulnerable areas such as rural North-West (89%) suggests that many mothers choose or are compelled to give birth at home due to constrained options. These constraints reflect broader systemic issues, including inadequate infrastructure, limited availability of skilled personnel, entrenched insecurity, and socio-cultural norms [50]. The role of inadequately equipped or staffed health facilities cannot be overlooked, with evidence indicating that fewer than 20% of healthcare facilities in Nigeria are capable of providing emergency obstetric care [51]. This observation is revealing, given that utilisation of services alone does not guarantee improved maternal or perinatal outcomes [52]. The global shift towards prioritising the quality of care calls for sustained, context-specific interventions in Nigeria [14, 52]. This premise also supports the urgent need for a social justice approach—one that goes beyond individual-level interventions to confront systemic barriers.

An important observation in our findings is that the South-East region deviated from the typical rural–urban pattern, with home birth prevalence slightly higher in urban areas (19%) than in rural areas (16%). Although the difference was marginal at the point estimate level, the finding may suggest a relatively uniform prevalence across settings in a region that also recorded the lowest national home birth (tied with the South-West). Current finding aligns with results from a previous study [19]. One plausible explanation lies in the unique socio-geographic characteristics of the South-East geo-political zone in Nigeria. Evidence supports a form of rural–urban symbiosis in the region, characterised by dense road networks, strong kinship ties, and frequent mobility [53], which may narrow the gap in infrastructure or access to services. Unlike more isolated or under-resourced rural areas elsewhere, rural communities in the South-East are said to benefit from communal development and diaspora investments [53]. Hence, rural residents in the region may have similar proximity to, and resources for utilising health facilities as their urban counterparts, potentially contributing to the parity observed.

Following multivariable analyses, home birth was associated with maternal and partner’s education, household wealth, ANC contacts, ethnicity, birth order, birth type, media and internet exposure, nationally and in our stratified analysis by urban-rural residence status. These findings align closely with evidence from studies in Nigeria [19–22, 41], other parts of Africa[44, 54], and globally [55]. While some of the factors identified might reflect individual decision-making, collectively, they are better understood as a reflection of broader social determinants of health and intersectionality, possibly shaped by structural inequities limiting equitable access to facility-based childbirth [32, 33, 56]. These interrelated factors could also reflect cumulative disadvantage. For instance, women with limited education, low household wealth, high parity, and poor access to ANC or health information could be systematically disadvantaged, not by choice, but by the structural conditions shaping their lives. Addressing these patterns requires context-specific, equity-focused policies that go beyond service provision to tackle root causes such as low education, poverty, and rural underdevelopment.

Specifically, maternal education was associated with home birth in a clear dose–response pattern. Women with no formal education, or only primary or secondary education, had increased odds of home birth than those with higher education, both in rural and urban areas. Education, or the lack of it, influences health-seeking behaviours and may reflect longstanding barriers to healthcare access, especially for women and girls in marginalised communities [57]. While education is a fundamental human right with empowering potential for making informed healthcare decisions [57, 58], Nigeria has the highest number of out-of-school children globally, with over 10 million school-age children, approximately 60% of whom are girls [59]. Nearly one in three primary school-age pupils and one in four junior secondary school-age children are not enrolled [59], highlighting a critical entry point for policy intervention. Partners’ education followed a similar pattern, reinforcing the potential influence of household dynamics and the importance of education. Where male partners have limited education, mothers may receive less support to seek skilled care.

Similarly, mothers from poorer households had significantly higher odds of home birth than their wealthier peers. This finding was evident in both rural and urban settings, underscoring the impact of economic inequality in Nigeria. The significantly higher odds of home birth among women in the middle wealth category in urban settings only, possibly reflect the reality of urban poverty, where families live in informal settlements or slums, underserved by health infrastructure, or the erosion of the middle class in urban Nigeria [60]. Furthermore, mothers with fewer than seven ANC contacts had a greater odds of home birth, with a more pronounced effect in rural areas. ANC contacts reflect both access to and engagement with the health system [61, 62]. Beyond clinical services, ANC can help build trust in the health system and support birth preparedness [62]. The pronounced rural effect of ANC suggests that structural factors, such as distance to health facilities, cost of services, or low-quality care, disproportionately affect rural women, reinforcing spatial health inequities.

Our analysis further reveals that mothers from the Hausa-Fulani ethnic group have significantly higher odds of home birth in both rural and urban Nigeria. This finding likely reflects a convergence of social and structural disadvantage. Many Hausa-Fulani communities, particularly those engaged in pastoralist and nomadic lifestyles, reside in geographically remote areas [63] with potential for limited access to schools and health infrastructure. These communities may also face compounded barriers, including cultural norms that may deprioritise facility-based care. The finding, thus, highlights how intersecting factors—ethnicity, mobility, geography, and socioeconomic status—can combine to restrict healthcare access or use. Other factors, such as higher birth order, multiple births, and low media exposure, highlight cultural, medical, and informational underpinnings. Higher-parity mothers may normalise home birth due to previous safe birth experiences, while multiple births may signal a higher risk of complications, necessitating health facility use. Conversely, limited media access can hinder exposure to vital health messages.

Notably, mothers who did not use the internet had higher odds of home birth, both in rural and urban areas. Internet access is recognised as a ‘super social determinant of health’, with the capacity to influence other factors such as employment, education and healthcare access [64, 65]. Limited internet use often reflects broader inequities [66], including educational and income constraints, which can restrict mothers’ ability to make informed care decisions. In rural areas, where healthcare access is presumably already limited, the lack of internet connectivity could further marginalise mothers. In urban areas, the digital divide persists, suggesting that economic and social inequities prevent even those in supposedly more developed settings from fully utilising digital resources. This finding supports the need for policies that promote digital inclusion. Integrating internet access into maternal health interventions can enhance equity in healthcare access [66] with potential for reducing the odds of home birth in Nigeria.

To understand within-population differences, we stratified our multivariable analyses by rural and urban residences. Findings remained largely consistent with those of an earlier study using a similar analytical approach [19]. The analyses revealed context-specific associations that align with broader social determinants of health and point to structural inequities. For example, the pronounced higher odds of home birth in rural settings across the North-East, North-Central, North-West, and South-South regions suggest persistent geographical inequities. These disparities are not merely regional; they potentially reflect how rurality amplifies disadvantage through poor infrastructure, workforce shortages, and limited institutional presence. The markedly higher odds of home birth in the rural

North-West and South-South may also exemplify how rurality and regional deprivation intersect to perpetuate maternal health disparities. From a social justice perspective, these patterns reflect historical underinvestment, sociopolitical marginalisation, and geographic inaccessibility.

In addition to the wide-ranging and pronounced regional differences, rural mothers also face distinct access-related barriers rooted in gender and power dynamics. For instance, difficulties in obtaining permission to seek care and the challenge of physical distance point to limitations on maternal autonomy and mobility. These barriers reflect both social norms and infrastructural deficits [34]. Our findings further indicate that mothers involved in healthcare decisions—either alone or jointly with partners—were less likely to give birth at home in rural areas. The association was not significant in urban areas, suggesting that a woman’s ability to make health decisions has a greater impact in rural regions. This finding supports interventions that empower women—both individually and in partnership with their spouses—to make informed decisions about their health [67]. Efforts to reduce home births and improve maternal health equity should include challenging restrictive gender norms and implementing husband/partner-inclusive interventions [67].

In urban areas, different factors came into focus. Financial barriers (paying for healthcare services) remained significant, despite the expected closer proximity to health services, suggesting that economic exclusion remains a key barrier in urban Nigeria. This pattern is consistent with the broader wealth index findings and is further supported by the significant association between home birth and middle wealth status in urban areas—a relationship not observed in rural settings. The findings indicate that in urban Nigeria, economic barriers to facility births impact not only the poorest but also the middle class. Contributing factors may include rising costs, a shrinking middle class, increased privatisation of healthcare services, and fewer alternative birth options, like traditional birth attendants, compared to rural areas. [13, 68]. Religion was also a significant factor in urban areas, with Muslim mothers having increased odds of home births than Christians or followers of other religions. This finding likely reflects misalignments between institutional care and religious or cultural expectations, highlighting the need for culturally sensitive maternal health services. Lastly, higher odds of home birth among younger urban mothers may reflect age-related vulnerabilities such as limited reproductive health knowledge, social support, or stigma [41]. These intersecting vulnerabilities require targeted policy responses to prevent early pregnancy and ensure adolescent and young adult mothers receive supportive, youth-friendly maternal care [41].

### Policy and programmatic implications

Our study highlights the need for targeted policies to address home births and improve maternal health outcomes in Nigeria. Prioritising policies that strengthen female education, especially in rural and northern areas, is essential. Expanding access to quality education for girls is a critical first step. Improving health literacy among mothers is also important, as it can help them understand birth complications and make informed healthcare decisions. Second, expanding access to communication infrastructure, nationally, should be a focus of policy efforts to facilitate the dissemination of culturally appropriate health messages and encourage community shifts toward skilled birth attendance. Third, infrastructure improvements are urgently needed to address geographic barriers. Investments in rural transportation networks, road infrastructure, and the construction of healthcare centres in northern and South-South regions are critical. Mobile health services and functional ambulance systems are vital for reaching remote populations and transferring patients to regional health centres. Developing the health workforce and ensuring a fair distribution of skilled birth attendants are essential. Improving access alone is not enough; there must also be a focus on enhancing the quality of care, particularly in life-saving emergency obstetric services[14, 52]. Fourth, targeted financial interventions are essential to overcome economic barriers in both rural and urban areas. In rural regions, poverty increases the likelihood of home births, while urban middle-income women also face similar trends. Policies should enhance subsidies for maternal services to improve affordability and equitable access.

Fifth, promoting maternal autonomy is vital, especially in rural areas. Engaging male partners and community networks in maternal health education can create supportive environments [67]. Programs that challenge gender norms and encourage shared decision-making are crucial for improving maternal health outcomes. Finally, our findings call for regionally tailored, culturally sensitive strategies that address the distinct patterns of inequality across Nigeria. In rural areas, policies should prioritise physical and informational access, women’s empowerment [67], and targeted investments in regions, particularly the North, where the Hausa-Fulani ethnic group is largely concentrated. In urban settings, the focus should be on improving financial accessibility, ensuring culturally sensitive care, especially for Muslim women, and addressing the specific vulnerabilities of younger mothers through youth-friendly services. Future research should assess the implementation, scalability, and effectiveness of these place-based strategies. In addition, further studies are needed to explore the role of cultural norms in shaping home birth practices in both rural and urban settings in Nigeria.

### Strengths and limitations

This study demonstrates key strengths, including analysis at national, urban, and rural levels, which clarifies disparities in home birth practice in Nigeria. The findings offer evidence-based insights for policies to reduce home births in Nigeria and are generalizable to the larger population due to the nationally representative sample. However, there are limitations to consider. Firstly, data collection relied on mothers’ recall ability, while it would be minimal given the substantial nature of the event, there appears to be a possibility of recall bias. Secondly, the study did not include potential culture-related factors that could influence home births in Nigeria. However, we captured wide-ranging factors based on the NDHS dataset, providing comprehensive insights. The dataset used is over five years old and may not accurately reflect the current situation. Nonetheless, it serves as a valuable starting point for further research on home births in Nigeria and other low- to middle-income countries. Finally, the data utilised were cross-sectional; hence, causality cannot be established, limiting the scope of our conclusions.

## Conclusion

Home birth remains widespread in Nigeria, significantly in rural areas and across all northern and South-South regions. Addressing home birth in Nigeria, therefore, requires both universal and targeted strategies. Interventions need to address the shared factors, such as improving education, expanding antenatal care access, and reducing financial and informational barriers, while also addressing the unique challenges within specific contexts. Rural areas require greater emphasis on physical access, women’s autonomy, and infrastructural investment, while urban strategies should consider economic vulnerability, youth-specific needs, and religious or cultural influences. Guided by a social justice lens, responses must be equity-oriented and locally grounded to ensure that all mothers, regardless of location, have access to safe, respectful, and high-quality maternal healthcare.

## Data Availability

The dataset can be accessed (https://dhsprogram.com/data/available-datasets.cfm) upon approval from the Demographic and Health Survey (DHS) Program.

## Acknowledgements

We are grateful to the DHS Program for providing access to the dataset used in this study. We also extend our sincere thanks to the mothers who participated, generously offering their time and valuable information. Additionally, we appreciate Oladimeji John Adewuyi for helping with preparing some of the Figs in this study.

